# Use of dried blood spot samples for SARS-CoV-2 antibody detection using the Roche Elecsys ® high throughput immunoassay

**DOI:** 10.1101/2020.10.19.20215228

**Authors:** Ranya Mulchandani, Ben Brown, Tim Brooks, Amanda Semper, Nicholas Machin, Ezra Linley, Ray Borrow, EDSAB-HOME Study Investigators, David Wyllie

## Abstract

**Background:** Dried blood spot samples (DBS) provide an alternative sample type to venous blood samples for antibody testing. DBS are used by NHS for diagnosing HCV and by PHE for large scale HIV and Hepatitis C serosurveillance; the applicability of DBS based approaches to SARS-CoV-2 antibody detection is uncertain.

**Objective:** To compare antibody detection in dried blood spot eluates using the Roche Elecsys ® immunoassay (index test) with antibody detection in paired plasma samples, using the same assay (reference test).

**Setting:** One Police and one Fire & Rescue facility in England.

**Participants:** 195 participants within a larger sample COVID-19 serodiagnostics study of keyworkers, EDSAB-HOME.

**Outcome Measures:** Sensitivity and specificity of DBS (the index test) relative to plasma (the reference test), at an experimental cut-off; quality of DBS sample collected; estimates of relative sensitivity of DBS vs. plasma immunoassay in a larger population.

**Results:** 18/195 (9.2%) participants tested positive using plasma samples. DBS sample quality varied markedly by phlebotomist, and low sample volume significantly reduced immunoassay signals. Using a cut-off of ten median absolute deviations above the immunoassay result with negative samples, sensitivity and specificity of DBS were 89.0% (95% CI 67.2, 96.9%) and 100.0% (95% CI 97.9, 100%) respectively compared with using plasma. The limit of detection for DBS is about 30 times higher than for plasma.

**Conclusion:** DBS use for SARS-CoV-2 serology, though feasible, is insensitive relative to immunoassays on plasma. Sample quality impacts on assay performance. Alternatives, including the collection of capillary blood samples, should be considered for screening programs.

## Introduction

Testing for SARS-CoV-2 antibodies is important to understand how the infection has spread in the population. However, extensive population testing is not feasible using the currently available, highly sensitive immunoassays, due to the need to take venous blood samples. As such, serosurveillance using home sampling is currently limited to the use of lateral flow immunoassays (LFIAs), which often have limited sensitivity^1^.

Dried blood spot (DBS) samples provide an alternative sample type to venous blood samples for antibody testing, and have been used extensively in screening for other viruses including Hepatitis B, Hepatitis C and HIV^2^. Recent studies have demonstrated the feasibility of using DBS for home blood collection for SARS-CoV-2 antibody screening, with or without virtual supervision^5^. Small scale feasibility studies have evaluated DBS samples for SARS-CoV-2 antibody detection in high risk populations, using plate based enzyme immunoassays, with promising results ^6,7^. Field studies indicate the DBS approach is likely to be acceptable^8^.

Recently, laboratories, including those in the United Kingdom (UK)’s National Health Service (NHS), have put in place high-throughput immunoassays for SARS-CoV-2 antibodies, which have to date used venous blood samples^9^. However, when considering population screening or individual risk assessment using antibody tests, an approach offering the potential advantages of easy and safe collection^10^ without the need for phlebotomy training and with the possibility for self-collection of samples^11^, stability of the sample at ambient temperature once dried^12^ and simple transport to the testing laboratory exempt from UN 3373 transport regulations^13^ would be attractive. The taking and transporting of DBS samples has proved successful in screening for other viruses^14^, and combined with testing on high throughput automated instrumentation could allow wider access to SARS-CoV-2 serology.

Here we describe a pilot study evaluating the potential for using DBS for SARS-CoV-2 antibody testing using the Roche Elecsys ® Anti-SARS-CoV-2 immunoassay. We chose this platform because it is widely available across the UK’s NHS, and has one of the best performance characteristics of existing assays, as evidenced by a large-scale study ^15^. The protocol used is an adaptation of that currently used by the PHE Manchester virology laboratory for high-throughput screening of Hepatitis C.

## Methods

### Participants studied

Samples were collected from volunteers attending two study locations over 2 days (4^th^ and 5^th^ June 2020): Site 1 - Lancashire Police Headquarters and Site 2 - Lancashire Fire and Rescue Service. This recruitment was part of a larger study of UK key workers, EDSAB-HOME, whose characteristics have been described elsewhere ^16^. Volunteers were key workers working on site during the pandemic, who were neither experiencing any COVID-19 compatible symptoms, currently or within the former seven. Eligibility was unaffected by any prior COVID-19 compatible symptoms or SARS-CoV-2 nasal and throat swabs, or antibody tests. The overall approach, study protocol, details of ethical approvals, and analysis for EDSAB-HOME are pre-specified in the study protocol, available at http://www.isrctn.com/ISRCTN56609224.

All EDSAB-HOME participants consented to provide a venous blood sample. All of those who attended a study clinic at Site 1 or 2 on the 4^th^ or 5^th^ June were asked whether they would like to provide an additional DBS sample; as such, they form a convenience sample. Sample collection was performed by 7 different phlebotomists, 5 at Site 1 and 2 at Site 2. The phlebotomists were experienced in the collection of venous blood samples but had no prior training in the collection of DBS samples, and a short on-site training session was provided.

### DBS collection system

High flow BD Microtainer® lancets were used for puncturing fingertip skin. We aimed to collect 4 full size spots onto a custom designed collection card. The card comprised PerkinElmer 226 grade collection paper and circle outlines for the collection of 5 separate 25μL blood spots, 4 of which are on a strip of paper attached with a perforated edge to enable removal using a disposable set of tweezers; each sample took ∼25-30 seconds to process. Samples were stored at room temperature in ziplock bags containing a dessicant pack, and returned to PHE Manchester virology laboratory overnight. Samples were processed for elution on the 11^th^ and 12^th^ June, and tested on 18^th^ June. Between elution and storage, they were stored at −80°C.

### Venous blood analysis (reference test)

Venous bloods were collected in 6ml volumes in EDTA Vacutainer® tubes (Beckton Dickinson) at the same time and sent to the PHE Seroepidemiology unit (SEU) Manchester. Plasma samples were separated from the EDTA blood after centrifugation at 1200 g for 15 minutes before sending to the PHE Rare and Imported Pathogens Laboratory (RIPL) Porton Down for testing. All plasma samples were tested with the Roche Elecsys ® Anti-SARS-CoV-2 immunoassay run on the Roche cobas e 801 analytical unit, following the manufacturer’s instructions (COI ≥ 1.0 was considered positive). Results from the venous blood analysis were treated as the reference standard in this study, chosen as this assay is currently the most sensitive and specific approach available for SARS-CoV-2 serological testing at PHE ^15^. Samples were sent from Public Health Laboratory Manchester on 17^th^ June 2020; due to limited availability of the Roche machines, samples were analysed at PHE Porton Down between 29^th^ June and 3^rd^ June (as part of a large batch of EDSAB-HOME study samples). Between receipt and analysis, all samples were stored at 4°C.

### Dried blood spot analysis (index test)

The size of the 4 spots collected on the card were assessed and recorded. A full-size spot filling the entire collection circle represents a DBS spot of 25μl in size. Samples were rated as “good” if 4 full size spots were collected on the card, see Figure 2a. Spots were considered “small” if they filled only 50% or more of the collection area, and “very small” if they filled less than 50%, see Figure 2b. A paper strip containing the 4 blood spots, the edges of which are pre-perforated, was removed using single use sterile forceps and placed in a 50ml skirted centrifuge tube (Eppendorff) containing 1 mL of PBS/Tween 0.05% (Sigma)

**Figure 1:**
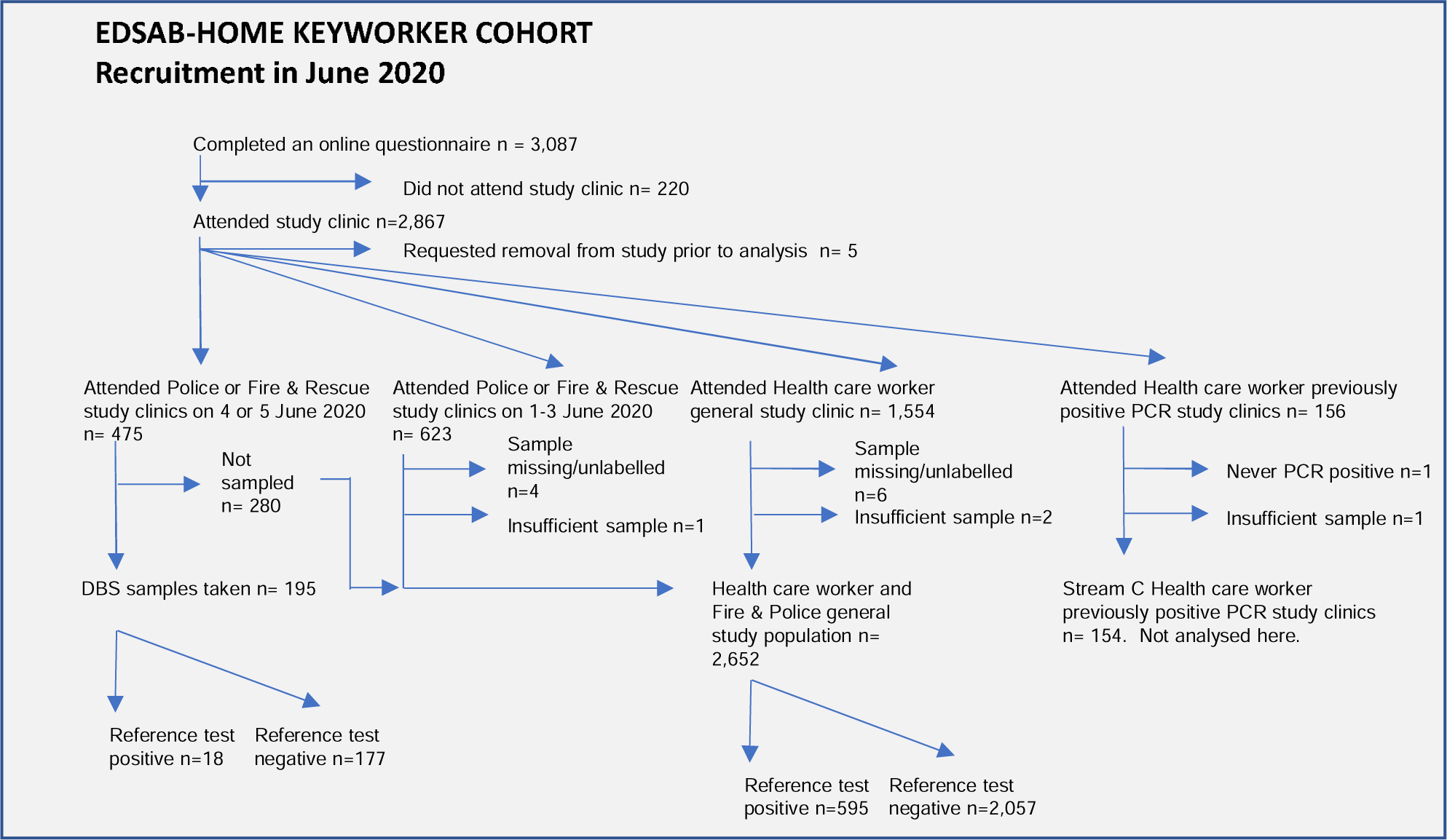
Flow diagram. Flow diagram illustrating where DBS samples were taken within the EDSAB-HOME study.

**Figure 2:**
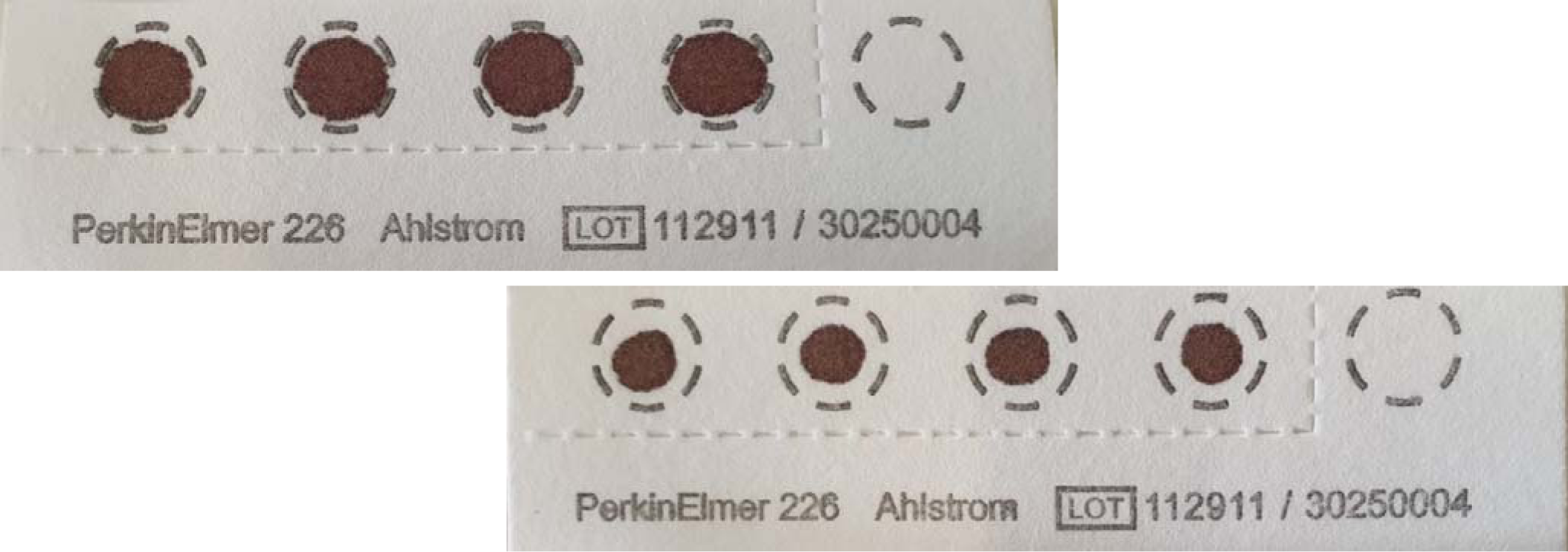
Dried blood spot collection kits. Images of dried blood spot collected on custom Perkin Elmer 226 collection card. The 4 blood spots are removed from the collection card using disposable tweezers and eluted into a single eluate. (a) 25μl dried blood spot: rated as “good”. DBS collected with spots smaller than those shown in the image were rated as small. (b) Image of “very small” dried blood spots collected on custom Perkin Elmer 226 collection card.

elution buffer. Tubes were placed flat on an orbital shaker for overnight elution at room temperature. Eluates were aspirated and transferred to false bottom tubes (Roche Diagnostics) and 12 μL tested with the Roche Elecsys ® Anti-SARS-CoV-2 antibody immunoassay run on the Roche e801 analyzer. Apart from using eluate instead of plasma, the protocol followed that was recommended by the manufacturer.

All reference tests were performed before the index tests. Those performing the index tests were blind to the reference test results. No clinical information on the samples was available to those performing either the index or reference test.

### Statistical Analysis

We described the quality of DBS samples overall, and by each individual phlebotomist. Quantitative results from index and reference assays were depicted graphically. Among cases positive on the reference test, linear models were fitted using the R *glm* function modelling log_10_(y) ∼ log_10_(x) + c + error, where y is the Roche immunoassay signal in the DBS eluate, x is the Roche immunoassay signal in the paired plasma sample, and c is a categorical variable representing spot size, which is either 0 (if the spot size is ‘good’) vs. 1 if it is small or very small.

For the index test, we derived an experimental cut-off, which is ten median absolute deviations about the median in samples negative by the reference test. An estimated limit of detection was computed from the regression model above as the most likely reference test value at which the index test values were at the experimental cut-off.

Sensitivity and specificity were computed by comparing the results of the dichotomised index test, using the experimental cut-off, relative to the dichotomised reference test result, using the manufacturer’s cut-off. All computations used R version 4.02. Receiver operating characteristic (ROC) analysis was performed using the pROC^17^ package, linear modelling used the *glm* function, and confidence intervals around proportions were computed using Wilson’s method with the Hmisc package *binconf* function.

In a separate analysis, we examined the distribution of the reference test (Roche Elecsys ® assay on plasma) in 2,652 samples from the EDSAB-HOME study, computing the immunoassay cumulative frequency distribution, and comparing it with the predicted limit of detection of DBS samples.

## Results

### Participants

We planned to recruit 200 individuals with paired venous and DBS samples from participants on 03 and 04 June 2020. Of the 475 individuals attending EDSAB-HOME study on those dates, 195 samples were collected, of which 18 were positive using the reference assay (Figure 1). Reasons for not collecting samples included subject or phlebotomist electing not to do so (which was permitted under the protocol). The demographics of those providing DBS samples were similar to those who did not (Table 1).

**Table 1:** Characteristics of the individuals eligible for DBS sampling.

|  |  | **Sampled**<br>n=195 | **Not sampled**<br>n=280 |
| --- | --- | --- | --- |
| **Age** | 18 to 24 | 9 (4.6%) | 10 (3.6%) |
|  | 25 to 39 | 62 (32%) | 96 (34%) |
|  | 40 to 59 | 115 (59%) | 168 (60%) |
|  | 60+ | 9 (4.9%) | 6 (2.1%) |
| **Gender** | Male | 126 (64%) | 168 (60%) |
|  | Female | 69 (35%) | 112 (40%) |
| **Ethnicity** | White | 181 (93%) | 266 (95%) |
|  | Not white | 14 (7%) | 14 (5%) |
| **Occupation** | Fire & Rescue | 48 (25%) | 50 (18%) |
|  | Police | 84 (43%) | 125 (45%) |
|  | Other | 63 (32%) | 105 (38%) |
| **Serostatus using plasma immunoassay (reference)** | Negative | 177 (91%) | 252 (90%) |
|  | Positive | 18 (9%) | 28 (10%) |

### Quality of DBS samples

Overall, around two thirds of the DBS samples were rated as “good” (n = 121, 62.1%); 42 (21.6%) were “small” and 32 (16.3%) were “very small” (Figure 2). The quality of sample collected varied by phlebotomist collecting the samples (Figure 3) ranging from 27/27 samples rates as “good” size to 0/21 rated as a “good” size.

**Figure 3:**
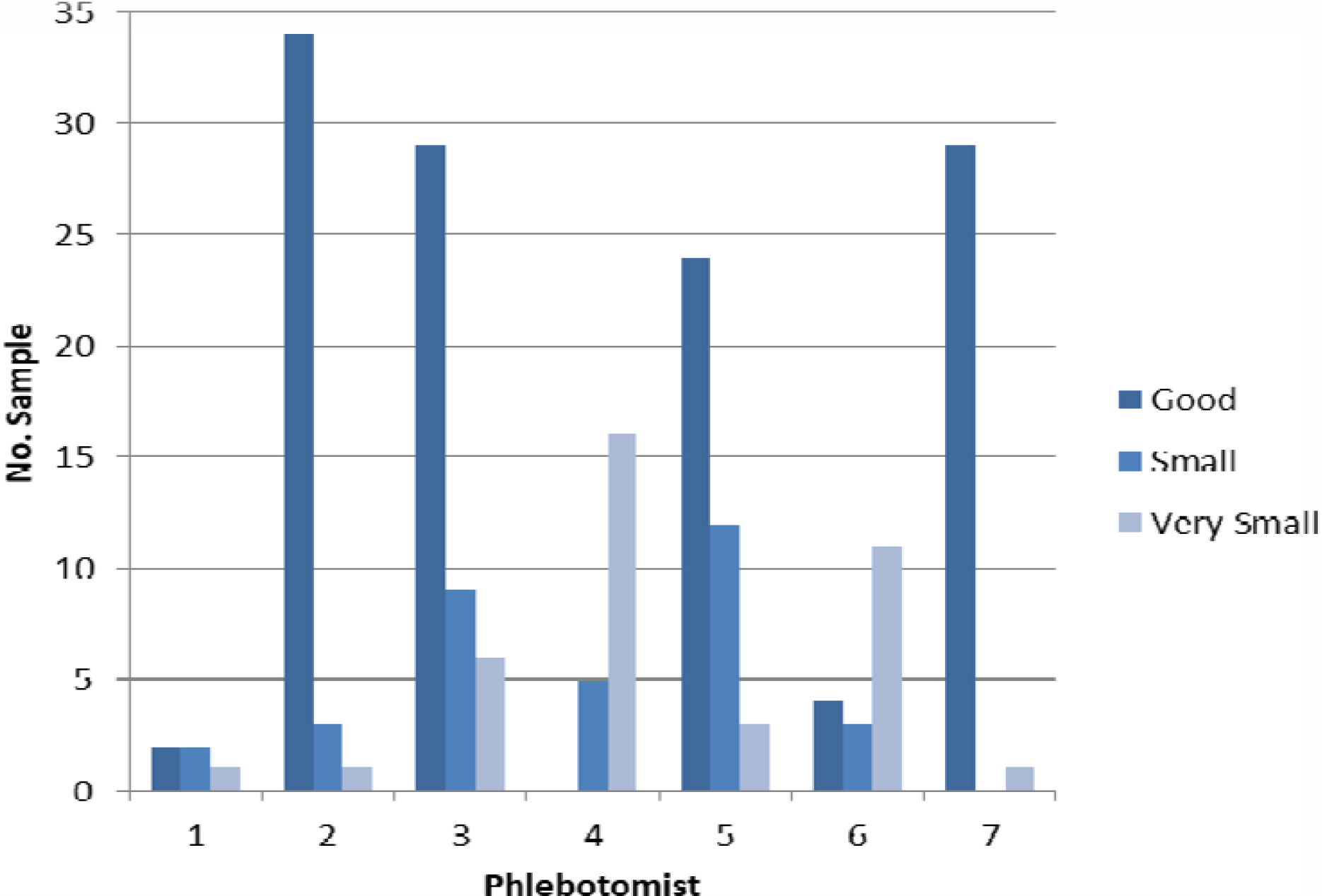
Quality of DBS sample collected by each phlebotomist. The quality scores of blood spots achieved by each of seven phlebotomists.

### Accuracy of the test applied to DBS samples relative to plasma samples

When we applied the manufacturer’s cutoff value, optimised for plasma, to the DBS eluates, the sensitivity relative to plasma was poor (44%, 95% CI 24.5, 66.2) (Table 2). We therefore considered whether the cutoff used for plasma was appropriate for our DBS eluates.

**Table 2.** Sensitivity and Specificity of index relative to reference immunoasssays. DBS samples compared to matched plasma samples tested on the Roche Elecsys ® Anti-SARSCoV-2 immunassay.

|  |  |  | **Venous plasma samples** |  | **Sensitivity % (95% CI)** | **Specificity % (95% CI)** |
| --- | --- | --- | --- | --- | --- | --- |
|  |  |  | Pos | Neg |  |  |
| **Manufacturer Positive Cut-off $\geq 1.0$** | Pos |  | 8 | 0 | 44.4<br>(24.5, 66.2) | 100<br>(97.9, 100) |
|  | Neg |  | 10 | 177 |  |  |
| **Experimental Positive Cut-off $\geq 0.17$** | Pos |  | 18 | 0 | 89.0<br>(67.2, 96.9) | 100<br>(97.9, 100) |
|  | Neg |  | 2 | 177 |  |  |

The relationship between the immunoassay ratio from venous plasma vs. DBS is shown for the 195 samples in Figure 4A. Although samples with negative reference tests gave very similar immunoassay ratios in both index and reference tests (black dots), positive samples generated lower results (median 0.7 vs. 18 units). Linear modelling showed that among positive samples, for every unit increase in antibody ratio in the reference test, index test ratio increased by a similar amount (Table 3). However, the index test (DBS) is much less sensitive: the limit of detection of the index (DBS) test was estimated as corresponding to 5.9 antibody index units in tests on plasma (Figure 4A). By contrast, the comparable figure limit of detection for the reference test, computed in the same way as the cut-off for the index test, and is 0.17, or about 33 fold lower. It is important to note that this estimate of limit of detection is based on “good” quality spots. Since index test ratios were 45% (95% CI 23, 82%) smaller in those with smaller spots *vs*. the 62% of “good” spots (Table 3), the test will be even less sensitive if smaller spots are present.

**Table 3:** Relationship between Roche immunoassay index using plasma and DBS. A general linear model was used to the DBS eluate Roche immunoassay result as a function of the plasma immunoassay result.

| Coefficient | Coefficient (95% CI) |
| --- | --- |
| Increase in $\log_{10}$(Roche DBS eluate) per unit increase in $\log_{10}$(Roche index on Plasma) | 1.09 (0.85, 1.31) |
| Fold change in Roche DBS eluate if sample is small or very small | 0.45 (0.23, 0.82) |

**Figure 4:**
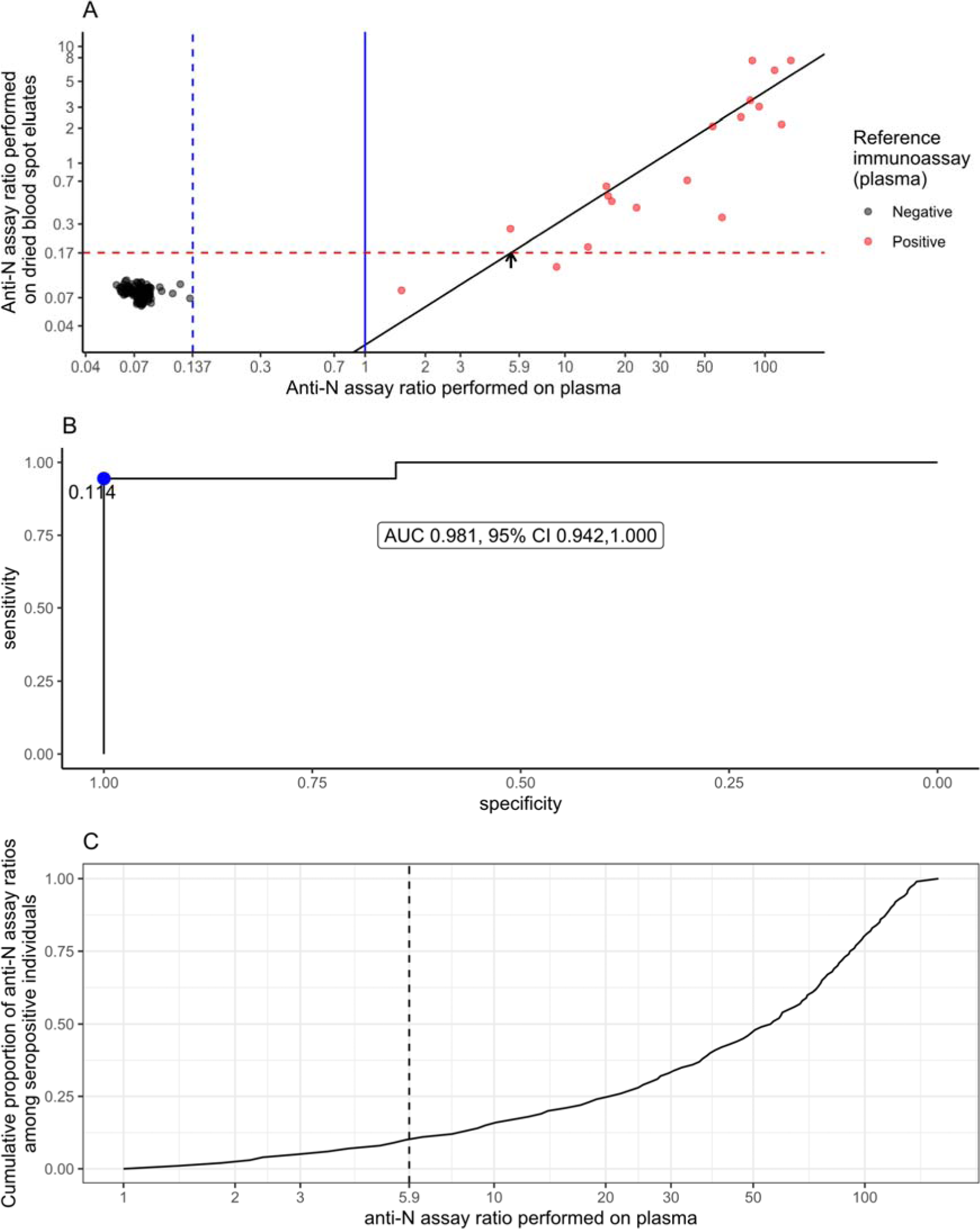
Immunoassay signals from plasma and from DBS samples. (A) The relationship between antibody ratios obtained using Roche Elecsys assays for plasma (reference text, x-axis) vs. paired dried blood spots (index test, y-axis). Samples positive in the plasma immunoassay are in red. Solid blue line: manufacturer’s cutoff for plasma samples (1). Red dotted line: experimental cutoff ten median absolute deviations above the reference test negative median for DBS (index test) samples (n=177). Blue dotted line: experimental cutoff ten median absolute deviations above the reference test negative median for plasma (reference test) samples. Solid black line: regression of index test immunoassay ratios on reference test immunoassay ratios. Arrow: estimate of limit of detection. (B) Receiver operator curve, showing sensitivity/specificity of DBS usage, relative to an immunoassay on plasma reference standard. (C) cumulative proportion of 440 seropositive cases from Police, Fire and Healthcare workers collected by EDSAB-HOME. There is no overlap with cases included in panel A. The estimated limit of sensitivity of the index test is shown as a dotted line.

Nevertheless, in this data set, the high correlation between tests results (Fig. 4A), and the small number of test results with low reference test results, and a predominance of “good” size spots, generates a high area-under-curve in ROC analysis (c= 0.985, Fig. 4B). Using the experimental cut-off derived above, of 18 positive samples using the reference test, 16 were positive in the index test giving a sensitivity estimate of 89.0% (95% CI 67.2, 96.9%), while maintaining a specificity of 100% (95% CI 97.9, 100%) (Table 2).

To understand whether this sensitivity is likely to be applicable to other samples, we examined the reference test (Roche Elecsys ® assay on plasma) in 2,652 samples from the EDSAB-HOME study (Fig. 1); there is no overlap between these samples and those on whom DBS samples were taken. Of these, 595 (22%) were positive on the reference assay, of which 62/595 (10.4%) were below the predicted limit of detection of DBS assays (Fig. 4C).

## Discussion

In this study, we showed that the DBS based approach used is an insensitive approach relative to assays on venous plasma, with the limit of detection of the assay estimated to be about 30 times lower with DBS samples. This is not unexpected, because the DBS eluate is at best 10% plasma. In reality, elution will be incomplete, and (as we showed) smaller spots result in lower concentrations of antibody. Nevertheless, in the small sample tested, the sensitivity of DBS at our experimental cut-off relative to the reference test was 89%, with wide confidence intervals due to the small study size. This is compatible with an observation, in a much larger independent cohort of 595 seropositive individuals, that about 10% of the population studied have antibody concentrations which we estimate would not be detectable by DBS.

This study has several important limitations. Firstly, DBS has only been tested on a relatively small sample set, only 18 of which were positive on the reference standard, and none of whom had a previous positive PCR test. Secondly, sensitivity and specificity estimates of the index test were based on cut-offs derived from the data, and so should be considered exploratory. Thirdly, the sensitivity of the test depends on the distribution of antibody concentrations in the target population; here, we only studied one population of key workers. If other cohorts are studied in which antibody concentrations have declined compared with the one we studied, DBS will perform worse than we observe here. Fourthly, we assessed samples taken by trained phlebotomist. Variation in performance was observed between phlebotomists (and presumably would also occur between self-taken samples); as such, usability studies to quantify the extent of this variation would be essential to understand how the tests perform when self-administered by the target population.

The key problem we have identified is lack of sensitivity. However, in populations with large amounts of antibody, such as individuals with more severe disease, or when very sensitivity assays are used, the different limits of detection of DBS vs. plasma samples may not translate into lower sensitivity. This may have occurred in the case of Morley *et al*, who report DBS performance to be comparable to matched serum samples, with a sensitivity of 98.1% and specificity of 100%, when compared to an in-house ELISA which detects Spike glycoprotein antibodies (using 87 samples from 80 volunteers, 37 who had had a previous PCR positive result)^18^.

When considering large-scale DBS testing programmes, additional factors which might impact on programme success include the more complex processing of DBS samples relative to plasma samples upon arrival in the testing laboratory. Nevertheless, this work indicates that for serosurveillance studies using the Roche Elecsys ® Nucleoprotein antibody assay, DBS based SARS-CoV-2 immunoassay workflows are feasible, subject to re-adjustment of cut-offs (as done in this study) and at the expense of sensitivity.

## Data Availability

Data can be made available on reasonable request via Public Health England's Office of Data Release.

## Acknowledgments

The authors would like to thank key collaborators: Chief Inspector Julie Rawsthorne from the National Police Wellbeing Service and Simon Fryer, Area Manager, Lancashire Fire & Rescue Service for their support in running clinics on site. We would like to thank Megan Lamb and Melissa Murove for their on-site support. We would like to thank all the phlebotomists, including Mo Phlebotomy, who took the venous and DBS samples. Finally, we would like to thank all those who volunteered to take part in this study.

## Sources of funding and role of funders

Public Health England.

## Conflicts of interest

EL & RB perform contract research on behalf of PHE for GSK, Pfizer and Sanofi Pasteur.

## Author contributions

Conceptualization; - RM,

Formal analysis; - RM, BB, DW

Investigation; - RM, DW, EDSAB-HOME study investigators

Methodology; - EDSAB-HOME study investigators

Supervision; DW

Validation; - DW, RM, BB.

Visualization; - DW

Roles/Writing - original draft; - RM, BB, DW

Writing - review & editing – RM, BB, TB, AS, NM, EL, RB, DW.

## Notes

### Competing Interest Statement

EL & RB perform contract research on behalf of PHE for GSK, Pfizer and Sanofi Pasteur. No other authors have any conflicts of interest to declare.

### Clinical Protocols

http://www.isrctn.com/ISRCTN56609224.

### Author Declarations

EDSAB-HOME study was approved by NHS Research Ethics Committee (Health Research Authority, IRAS 284980) on 02/06/2020 and PHE Research Ethics and Governance Group (REGG, NR0198) on 21/05/2020.

